# Examining Australian’s beliefs, misconceptions, and sources of information for COVID-19: A national online survey

**DOI:** 10.1101/2020.07.27.20163204

**Authors:** Rae Thomas, Hannah Greenwood, Zoe A Michaleff, Eman Abukmail, Tammy Hoffmann, Kirsten McCaffery, Leah Hardiman, Paul Glasziou

## Abstract

**Objective:** Public cooperation to practice preventive health behaviours is essential to manage the transmission of infectious diseases such as COVID-19. We aimed to investigate beliefs about COVID-19 diagnosis, transmission and prevention that have the potential to impact the uptake of recommended public health strategies.

**Design:** An online cross-sectional survey conducted May 8 to May 11 2020.

**Participants:** A national sample of 1500 Australian adults with representative quotas for age and gender provided by online panel provider.

**Main outcome measure:** Proportion of participants with correct/incorrect knowledge of COVID-19 preventive behaviours and reasons for misconceptions.

**Results:** Of the 1802 potential participants contacted, 289 were excluded, 13 declined, and 1500 participated in the survey (response rate 83%). Most participants correctly identified “washing your hands regularly with soap and water” (92%) and “staying at least 1.5m away from others” (90%) could help prevent COVID-19. Over 40% (incorrectly) considered wearing gloves outside of the home would prevent them contracting COVID-19. Views about face masks were divided. Only 66% of participants correctly identified that “regular use of antibiotics” would not prevent COVID-19.

Most participants (90%) identified “fever, fatigue and cough” as indicators of COVID-19. However, 42% of participants thought that being unable to “hold your breath for 10 seconds without coughing” was an indicator of having the virus. The most frequently reported sources of COVID-19 information were commercial television channels (56%), the Australian Broadcasting Corporation (43%), and the Australian Government COVID-19 information app (31%).

**Conclusions:** Public messaging about hand hygiene and physical distancing to prevent transmission appear to have been effective. However, there are clear, identified barriers for many individuals that have the potential to impede uptake or maintenance of these behaviours in the long-term. Currently these non-drug interventions are our only effective strategy to combat this pandemic. Ensuring ongoing adherence to is critical.

**What is already known on this topic:**

- The current strategies to prevent the transmission of COVID-19 are behavioural (hand hygiene, physical distancing, quarantining and testing if symptomatic) and rely on the public knowledge and subsequent practice of these strategies.
- Previous research has demonstrated a good level of public knowledge of COVID-19 symptoms and preventive behaviours but a wide variation in practicing the recommended behaviours.
- Although knowledge can facilitate behaviour change, knowledge alone is insufficient to reliably change behaviour to the widespread extent require to combat health crises.

**What this study adds:**

- Participants reveal confusion about whether wearing masks will reduce transmission, apprehension about attending health services, and perceptions that antibiotics and alternative remedies (such as essential oils) prevent transmission.
- Analysis of why participants hold these beliefs revealed two dominant themes: an incomplete or inaccurate understanding of how COVID-19 is transmitted, and the belief that the behaviours were unnecessary.
- This study underlines the necessity to not only target public messaging at effective preventative behaviours, but enhance behaviour change by clearly explaining why each behaviour is important.

## Introduction

So far, Australia has done well in its response to the severe acute respiratory syndrome coronavirus 2, commonly called COVID-19, but not because of vaccines or drug treatments. The success is due to the use of non-drug interventions, such as physical distancing, hand hygiene, and quarantining [1]. Maintaining the suppression of major outbreaks are critical but it is predicated on garnering public cooperation to continue to conduct appropriate public health behaviours. Misinformation about prevention, transmission, and treatment of COVID-19 has the potential to derail these efforts.

To correct misinformation, we must first identify what individuals believe, which beliefs are accurate and inaccurate, and the reasons for inaccurate beliefs. Understanding the reasoning behind inaccurate beliefs and exploring typical sources of information can facilitate behaviour change interventions designed to support risk reduction efforts [2-4]. Several surveys have explored people’s perceptions, knowledge and attitudes of COVID-19. International and Australian-focused online surveys suggest 71% −93% of participants [5,6] could *identify* preventive behaviours (e.g., hand hygiene, physical distancing), but the proportion of individuals who self-report *practicing* these behaviours were fewer, ranging from a low of 45% in a survey of UK participants [7] to 85% in an Australian sample [8]. The difference between the high proportion of people *knowing* appropriate prevention behaviours and the lower proportion of those reporting to *practice* the behaviour, suggests that knowledge alone is insufficient to change behaviour. This constitutes a knowledge-practice gap. Varying demographic differences have been reported to predict practicing preventive behaviours including low socio-economic status [7], low health literacy [5], anxiety [5,8], and those with high compared with low engagement and perceptions of risk [8,9].

To achieve high compliance with infectious disease prevention measures relevant to the current COVID-19 pandemic and future infectious outbreaks, we need to understand the reasoning behind why some people do not practice effective prevention behaviours [2]. Therefore, we aimed to investigate beliefs about COVID-19 that have the potential to impact the uptake of the appropriate prevention behaviours (particularly the public health messages from the Australian Government), why some participants hold incorrect beliefs, and the most commonly used sources for acquiring COVID-19 related information.

## Methods

### Study design and participants

A cross-sectional, online survey of eligible adult Australians was conducted from May 8 to May 11 2020. The national sample, with representative quotas for age and gender, was provided by online panel provider, Dynata (https://www.dynata.com/). The online survey was scripted using Qualtrics (https://www.qualtrics.com/au/). Bond University Human Research Ethics Committee provided ethics approval for this research (#RT03008).

People aged 18 years and over and living in Australia were eligible to participate. Healthcare professional and people who had been tested for COVID-19 were ineligible to participate as their knowledge about COVID-19 may differ compared to the Australian public. Prior to commencing the survey, potential participants read a detailed study explanatory statement. Continuation of survey was accepted as informed consent.

### Patient and public involvement

A member of our research team and co-author (LH) is a consumer representative and provided advice on common misconceptions identified in her role as consumer advocate. These assisted in developing the search for misconceptions to inform the survey content. Other patient and public involvement was not sought.

### Survey

The survey was informed by two searches which aimed to identify public misconceptions that, if believed, would have the potential to negatively impact the uptake of the behaviours recommended by public health authorities. First, we conducted a focused literature search of PubMed for misconceptions from the current COVID-19 pandemic and previous epidemics (i.e., keyword search included terms for severe acute respiratory syndrome, H1N1 Influenza, Middle East respiratory syndrome coronavirus and myths or misconceptions). We also conducted an environmental scan of internet and social media sites (e.g., Facebook) using the terms “COVID-19” and “myths”. Finally, we conducted citations checks of identified articles (research publications and internet/news articles) to detect further misconceptions and sources of information. Misconceptions were categorized as relating to the diagnosis, transmission and prevention of COVID-19. The survey was piloted with colleagues and members of the public to ensure face and content validity, and ease of completion.

### Measures

The survey consisted of 5 sections (COVID-19 symptoms, prevention strategies, beliefs for ‘incorrect’ prevention strategies, transmission behaviours, and information sources for COVID-19). The survey items and response scales are available in the Supplementary file. The expected time to complete the survey was 10 minutes.

### Statistical Analysis

The primary outcome was the frequency of prevention strategy misconceptions and participant reasoning for each. Descriptive statistics (counts and percentages) were calculated for participant demographic variables and correct/incorrect survey responses. ‘Correct’ and ‘incorrect’ answers for survey questions were determined by the study team via group consensus a priori.

For the open response options, we conducted content analyses of the ‘incorrect’ responses. Two authors (RT and LH) independently coded the first 50 responses and developed an initial coding framework. Uninformative responses (e.g., “I don’t know”; “unsure”) were not coded. Codes were compared and the framework was iteratively developed until consensus was reached. The same two authors tested the refined the coding framework on the next 20 responses. Following consensus, one author (RT) complete the qualitative analyses for the remaining responses. Due to the large unexpected volume of ‘incorrect’ responses for three items (wearing masks; wearing gloves; and staying away from healthcare centres), a random 50% of the responses was coded. All responses were coded for the other four items (use of colloidal silver and essential oils, use of antibiotics, social distancing, washing hands).

## Results

Of the 1802 potential participants contacted, 289 were excluded, 13 declined, and 1500 participated in the survey (response rate 83%; see Supplementary Figure 1 for participant flow chart). As per the sampling frame, 50% of survey participants were female, there was a representation across adult age groups and a proportional representation from each Australian state and territory. Participant characteristics are reported in Table 1.

**Table 1.** Participant characteristics (N=1500)

| **Characteristics** | **N** | **%** |
| --- | --- | --- |
| Female | 750 | 50 |
| Age |  |  |
| 18-24 y | 171 | 11 |
| 25-34 y | 264 | 18 |
| 35-44 y | 239 | 16 |
| 45-54 y | 223 | 15 |
| 55-64 y | 222 | 15 |
| 65-74 y | 227 | 15 |
| 75 or older | 154 | 10 |
| Education |  |  |
| High school graduate or Less | 459 | 31 |
| Trade Certificate (I-IV) | 276 | 18 |
| Tertiary | 765 | 51 |
| Australian States and Territories |  |  |
| QLD | 302 | 20 |
| NSW | 471 | 31 |
| ACT | 29 | 2 |
| NT | 9 | 1 |
| WA | 160 | 11 |
| VIC | 382 | 25 |
| TAS | 34 | 2 |
| SA | 113 | 7 |
| Aboriginal or Torres Strait Islander |  |  |
| Yes | 17 | 1 |
| No | 1471 | 98 |
| Prefer not to say | 12 | 1 |
| Born in Australia | 1049 | 70 |

### Identifying prevention strategies

A high proportion of the sample gave correct responses for hand hygiene and 1.5m physical distancing, but significant proportions were incorrect for the effectiveness of antibiotics, colloidal silver, wearing gloves or masks, and staying away from health centres. Figure 2 shows proportions of participant perceptions of preventive behaviours and Table 2 reports the results of the content analyses for the ‘incorrect’ responses.

**Table 2.** Content analyses of misconceptions of possible preventive behaviours.

|  | Behaviour/belief | Major themes for non-compliance (%) | Quotes |
| --- | --- | --- | --- |
| Appropriate behaviours | Washing hands with soap regularly<br>(N=24; all coded) | Unnecessary or ineffective<br>(37%) | “because COVID is not only on your hands but can be on other parts of your body”<br>“because we are always touching different surfaces”<br>“it is an overreaction to wash hands too often” |
|  |  | Incorrect or incomplete understanding of transmission<br>(17%) | “it’s air born”<br>“because most people will catch the disease from breathing it in”<br>“the body needs germs to wash regularly damages the immune system” |
|  | Physical distancing (1.5 mtrs)<br>(N=34; all coded) | Unnecessary or ineffective<br>(50%) | “I don’t think that social distancing is really effective”; “if u [sic] are going to get it you’ll get it”<br>“It is crap, if someone is not sick, then keeping away or close makes no difference” |
|  |  | Incorrect or incomplete understanding of transmission<br>(12%) | “It doesn’t help anything if people have the disease”<br>“because it can jump from person to person and move through the air” |
| Inappropriate behaviours | Taking antibiotics to prevent COVID<br>(N=167; all coded) | Incorrect understanding of antibiotics and treatment for COVID-19<br>(30%) | “it helps to fight the virus”<br>“keep you strong”<br>“antibiotics kills any infection in your body and stops you from getting sick” |
|  |  | Strengthens immune system<br>(29%) | “continued boosting of the immune system and supporting it”<br>“increases your immune system” |
|  | Using products like colloidal silver or essential oils<br>(N=179; all coded) | Incorrect or incomplete understanding of transmission and treatment options for COVID-19<br>(32%) | “colloidal silver kills germs, fungus and bacteria...”<br>“.... it could burn out the infection”<br>“I understand that they dilute the virus” |
|  |  | Strengthens immune system<br>(9%) | “would build up your immune system”<br>“... giving your body the best chance to fight it” |
| Contentious behaviours | Wearing gloves away from home (N=596; random ½ coded; n=299) | Incorrect or incomplete understanding of transmission (33%) | “stop infection transmitting”<br>“pick up any germs from anything or anyone” |
|  |  | Plausible self-protection (18%) | “to stop me from contracting something and putting it on my face”<br>“helps prevent touching contaminated surfaces”<br>“they form a barrier to my touching virus-laden surfaces”. |
|  | Wearing a surgical mask when away from home (N=623; random ½ coded; n=314) | Incorrect or incomplete understanding of transmission (33%) | “to avoid contamination in my mouth”<br>“so you won’t get sick”<br>“it can prevent transmission” |
|  |  | Plausible self-protection (23%) | “it stops you from touching around your mouth”<br>“masks help reduce the risk of a person becoming infected by inhaling the virus”<br>“I think it would at least lessen the risk of becoming infected by covering two of the areas the virus can enter the body through” |
|  | Staying away from hospitals and health centres (N=872; random ½ coded; n=435) | Infected patients and staff (52%) | “hospitals are the source of the disease at the moment as hospital is designed for sick people and it is covid-19 era at the moment”<br>“Because hospitals and health centres are where other people who could be infected may be” |
|  |  | Explicit concerns of safety and risks (15%) | it helps to stay away from potential risk<br>Reduce risk of contact with infected people<br>Hospital is risky zone now |
|  |  | Negative beliefs of health services (14%) | those places are where most germs of any description are<br>Ideal places to pick up bugs. Go there only in emergency |

**Figure 1.**
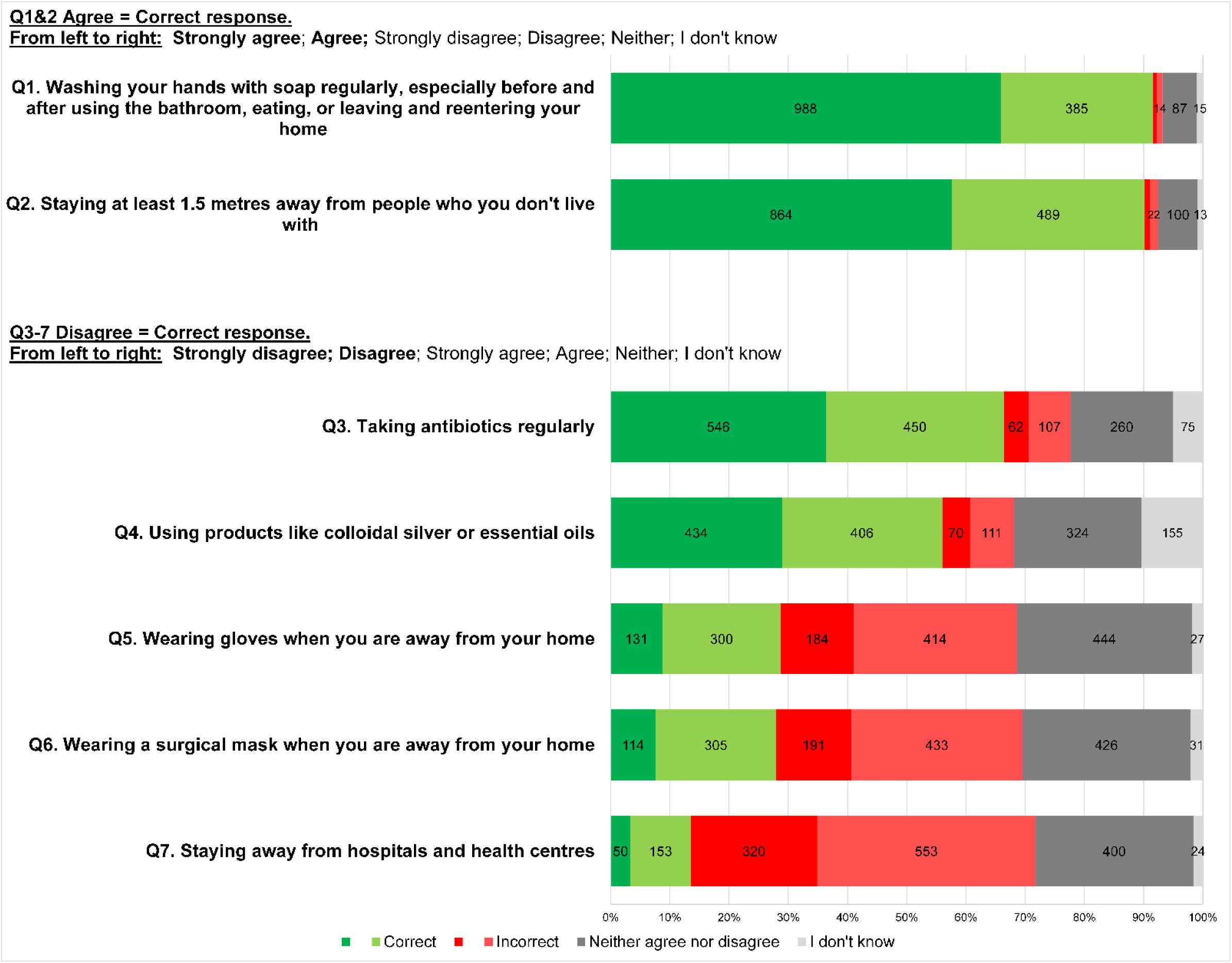
Participant perceptions of possible preventive behaviours.

**Figure 2.**
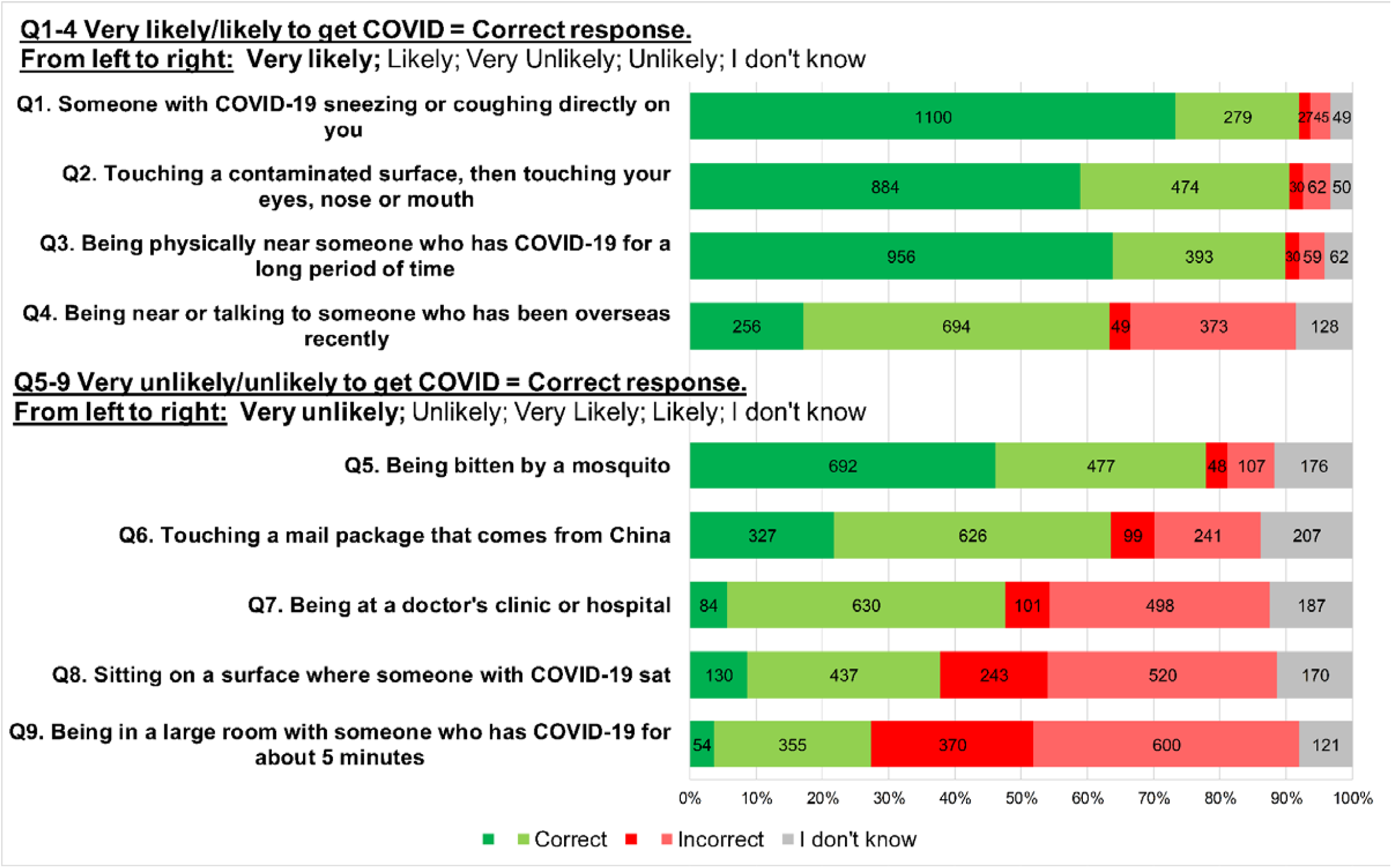
Participant perceptions of their likelihood to contract COVID-19.

Though most participants correctly identified “washing your hands regularly with soap and water” (92%), 7% were undecided (“neither agree or disagree” or “I don’t know”) and a minority (2%) thought that washing their hands would NOT prevent COVID-19. Reasons varied, but the most frequent theme (37.5%) was that participants thought handwashing was unnecessary or ineffective (e.g., it was an “overreaction”, it wouldn’t ‘kill’ the virus, or it is redundant because the participant didn’t leave the house; see Table 2).

Similarly, 90% of participants correctly identified that “staying at least 1.5m away from others” could prevent COVID-19. Only 34 (2%) participants thought that physical distancing would NOT prevent COVID-19 transmission. Half of those who held that belief reasoned that the strategy was ineffective or unnecessary (e.g., “stays in the air” and “can float a lot further than that”; see Table 2).

Fewer participants correctly identified that “regular use of antibiotics” (66%) would NOT prevent COVID-19, and 22% were unsure. For those who thought that antibiotics *could* prevent COVID-19 (11%), two dominant themes emerged: an incorrect understanding of antibiotics and treatment options for COVID-19 (e.g., they could “block viruses”, and “prevent” or “kill” the virus) and erroneously thinking antibiotics would strengthen their immunity.

While 56% of participants correctly identified that taking “products like colloidal silver or essential oils” would NOT prevent a person from contracting COVID-19 and 32% were unsure, and the remainder thought those products would help to prevent COVID-19. Approximately 32% of those who thought colloidal silver and essential oils would prevent COVID-19 appeared to incorrectly understand the transmission process and prevention strategies for COVID-19. For example, participants reasoned these products would help because they were “good in disinfecting”, suggesting transmission only by touch and “evidence that colloidal silver is a virus killer” and the products would “kill the germs”.

The two contentious preventative measures of wearing gloves or wearing surgical masks when outside of the home, showed conflicting beliefs about the benefits of these behaviours to prevent COVID-19. Over 40% of participants considered wearing gloves outside of the home would help prevent them contracting COVID-19. Their reasons were nuanced. In the randomly coded sample of 50% (n=299), 33% of participants incorrectly understood the transmission of COVID-19 (e.g., “won’t come into contact with the virus”, “stop transmission”). However, 23% identified plausible self-protection reasons (e.g., “would not be touching surfaces where it might be on and then touching my face and rubbing my eyes with bare hands”, “reduces risk of uptake of pathogen from hands, which in turn reduces risk of transmission from hands to eyes, nose, mouth, etc”).

For “wearing of face masks outside of the home”, 28% of participants agreed that this would NOT prevent them (the wearer) from contracting COVID-19, while 42% of participants thought this behaviour was preventative, and the remainder unsure. Within the randomly coded sample, the most frequently provided reason for wearing surgical masks (33%) was an incorrect understanding of COVID-19 transmission (e.g., “because the virus cannot move via a surgical mask”, “you can’t catch it”). Yet, almost a quarter (23%) identified plausible reasons for wearing masks (“may prevent inhaling droplets from the air which may contain the COVID-19 virus”, “it is some form of barrier to my respiratory system”).

Most participants agreed (58%) or were unsure (28%) that staying away from hospitals and health centres would prevent COVID-19 transmissions. Of the randomly coded responses, 52% articulated concerns of infection from test-positive COVID-19 patients and staff, while 13% had reasoned that hospitals and health centres should be avoided in general (e.g., “hospitals are hotbeds of disease”).

### Understanding transmission process

When asked to identify ways in which they were likely or unlikely to contract COVID-19, participant responses were again varied (Figure 3).

The majority of participants correctly identified that transmission of COVID-19 was likely to occur though “sneezing/coughing” (92%), “touching a contaminated surface and then touching your eyes, nose or mouth” (91%) and “being physically near someone with COVID-19 for a long period of time” (90%). Fewer participants (63%) correctly identified that “being near someone who had recently been overseas” made it more likely that they could contract COVID-19. Most participants also correctly identified that you are unlikely to contract COVID-19 from “mosquito bite” (78%) or by “touching a package that had come from China” (64%).

In contrast, 65% of participants thought it was very likely or likely they would contract COVID-19 due to “being in a large room with someone for a short period of time” and 51% thought they had a very likely or likely chance by “sitting on a surface where someone with COVID-19 sat”. In a similar response pattern to preventing transmission, there was a lot of uncertainty about whether “attending a doctor’s clinic or hospital” increased your risk of contracting COVID-19with an even proportion of participants thinking it was unlikely this would increase their risk (48%) and those who thought it would (40%) or were unsure (12%).

### Awareness of COVID-19 symptoms

The majority of participants (90%) identified “fever, fatigue and cough” as indicators of COVID-19 and which, if experienced, would require a person to take further precautions (Table 3). However, just over a third of participants (35%) correctly identified “fever, fatigue and cough” as the *only* indicator (Supplementary Table 2). Of concern, 42% of participants thought that being unable to “hold your breath for 10 seconds without coughing” was an indicator of having COVID-19 (Table 3).

**Table 3:** Frequency of incorrect symptom/ behaviour responses.

| **Response** | **N (%)** |
| --- | --- |
| You cannot hold your breath for 10 seconds without coughing | 625 (42) |
| You have a runny nose and wet cough | 503 (34) |
| You have had your temperature checked and it is normal | 234 (16) |
| You have a rash | 167 (11) |

Note: participants were able to choose more than 1 response.

### Information sources

The most frequently reported sources of COVID-19 information were Channels 7,9 10 news (56%), ABC news (43%), and the Australian Government COVID 19 app (31%). Supplementary Table 3 shows the frequency in which each source of information was ranked as first, second or third. Only 23% of participants reported social media as a top 3 source of information and of those did, less than a quarter (24%) of these ranked it as their first source of information.

## Discussion

We aimed to identify common beliefs, and describe reasons for inaccurate beliefs, about COVID-19. We specifically focused on beliefs or behaviours that have the potential to inhibit the uptake of the Australian public health recommendations for COVID-19 and those that might hamper the uptake of effective preventive behaviours in the future.

The widespread ability to identify the preventative measures of hand hygiene and physical distancing suggests that public health messaging has been effective in communicating prevention strategies to Australians. Only a minority of participants thought these behaviours were unnecessary or ineffective. Additionally, almost all participants correctly identified key public health messaging of common symptoms of fever, fatigue and cough and potential transmission practices such as sneezing and coughing and touching contaminated surfaces. This prevention awareness is supported by several other surveys with between 71% and 93% [5,6,8] of participants endorsing correct responses or reporting practicing the behaviour.

However, when we examined preventative behaviours not explicitly communicated by public health authorities, the results are more varied. While two-thirds of participants correctly identified that taking antibiotics would not prevent COVID-19, the remaining third thought taking them *would* prevent COVID-19 or were unsure. When asked for reasons for this misconception, it emerged that most who endorsed antibiotics had poor understanding of how they worked and their effects on viruses and the immune system broadly. Of concern, this result is similar to another Australian study that reported 35% of participants also thought antibiotics would be effective to prevent or treat COVID-19 [8]. Inappropriate use of antibiotics is considered the leading cause of antibiotic resistance [10,11]. The perception that antibiotics could prevent or treat COVID-19 has the potential to derail ongoing efforts to curb antibiotic use in the community [12]. Further, only half of participants correctly identified that products like colloidal silver or essential oils are *not* effective preventing transmission of COVID-19. Taken together, it appears many Australians have misconceptions about the effectiveness of pharmaceutical and complementary medicine’s roles in preventing COVID-19. If people believe these approaches will stop them from contracting the virus, there is legitimate concern they will not practice non-drug interventions such as hand hygiene and physical distancing either now or in the future.

Equally concerning was our finding that at the time of the survey (early May) only 15% of participants were confident that visiting hospitals and health centres were safe. Australian data from the Medical Benefits Scheme [13] appears to reflect this uncertainty with a 23% drop in face-to-face primary care attendance compared to the average of the same month in the past 5 years. A sharp decline in presentations to Emergency Departments for reasons other than COVID-19 (e.g., stroke, cardiac events etc) has also occurred internationally [14,15]. Conversely, the increase in telemedicine (e.g., video and phone consultations) in Australia [13] may suggest that although people are physically staying away from physically attending health services, they are still receiving health care through other means. These findings suggest that the emergence of COVID-19 has resulted in significant changes in how healthcare is utilised. The impact of these changes in term of accessing preventative healthcare services, patient’s health outcomes and healthcare costs are yet to be determined [16].

Two preventative behaviours that were associated with a high degree of uncertainty were wearing gloves and wearing a mask outside of the home. Approximately 40% of participants believed wearing gloves or a mask would prevent transmission of COVID-19, similar to Seale et al’s findings [8] that 50% of surveyed Australians thought their use could prevent COVID-19. This uncertainty surrounding wearing a mask is likely a reflection of the mixed messages that are evident in the media and various public health campaigns used in both Australia and internationally. These inconsistent and unclear messages may have contributed to the misconceptions about the effectiveness of masks and how they can reduce the transmission of COVID-19. There has been no public health messages in Australia suggesting wearing gloves is an effective preventive behaviour. In both cases, many participants did not fully understand how transmission of COVID-19 occurred. With many thinking that either masks or gloves would be sufficient to stop the spread. This is the primary concern of public health authorities; that should people wear masks, they may not practice more effective preventive behaviours such as hand hygiene and physical distancing because they believe masks and gloves are enough to protect them from COVID-19. However, some recognised the additional benefit that masks might provide.

Overall, our findings are concordant to those of a recent online survey of over 4000 Australians which also found gaps in understanding. This study found that those with inadequate health literacy and who spoke a language other than english were less able to identify behaviours to prevent infection and experienced more difficulty finding information and understanding public health messaging about COVID-19 [5]. In McCaffery et al [5], there were also higher endorsements of misinformation including negative views of vaccines amongst the same groups. Given that these views have broader public health implications (decreased vaccine uptake, increased antibiotic resistance), it is critical that we target future messaging to disadvantaged populations such as those with low health literacy and those who are culturally and linguistically diverse to ensure that health consumers are better equipped to make informed decisions with regards to their health care.

## Strengths and limitations

In our sample, 70% of participants were born in Australia. Although this reflects Australian population demographics, results from this survey may not translate to individuals where English is not their primary language. We did not assess cultural and linguistic diversity which limits the generalisability of our findings to this group. McCaffery et al [5] identified that participants whose primary language was not English had poorer understanding of COVID-19 symptoms, were less able to identify behaviours that reduced infection risk, and experienced more difficulty understanding Government messaging. Public health messaging not only should ensure translation of messages but also understand the cultural differences that may interfere with practicing preventive behaviours. Unlike other Australian surveys [5,8], our survey looked beyond quantifying knowledge and attitudes to include qualitative content analyses to identify specific reasons for participants’ misconceptions.

Our sample was balanced for gender and stratified for age. We also had proportionally representative quotas from each Australian State and Territory. Despite these efforts, we acknowledge that survey research (with panel providers) has certain biases such as selection and sampling bias, and therefore suggest care when interpreting any results.

## Conclusion

Our results suggest that although clear public messaging about the two, key evidence-based prevention behaviours (hand hygiene and physical distancing) are broadly understood, there are still important knowledge gaps around how the disease is prevented, transmitted and its symptoms. Public health messages will need to combat the misconceptions that antibiotics and complementary medicines are effective in prevention and/or treating COVID-19. These beliefs have the potential to impede individuals from practicing appropriate and effective behaviours. In the absence of a vaccine or effective drug treatments our only prevention strategies are non-drug interventions, especially physical distancing and hand hygiene [1]. How do we maintain these behaviours in the long term and how do we initiate other behaviours yet to be seen as critical such as wearing masks, testing when symptomatic, self-isolating, and downloading contact tracing apps? By understanding some of the misconceptions identified in our survey and using the principles of public communication and knowledge translation, we can develop intervention strategies for the longer term.

## Data Availability

Data will be available upon request once manuscript is published.

## Supplementary file

**Supplementary Table 1.**
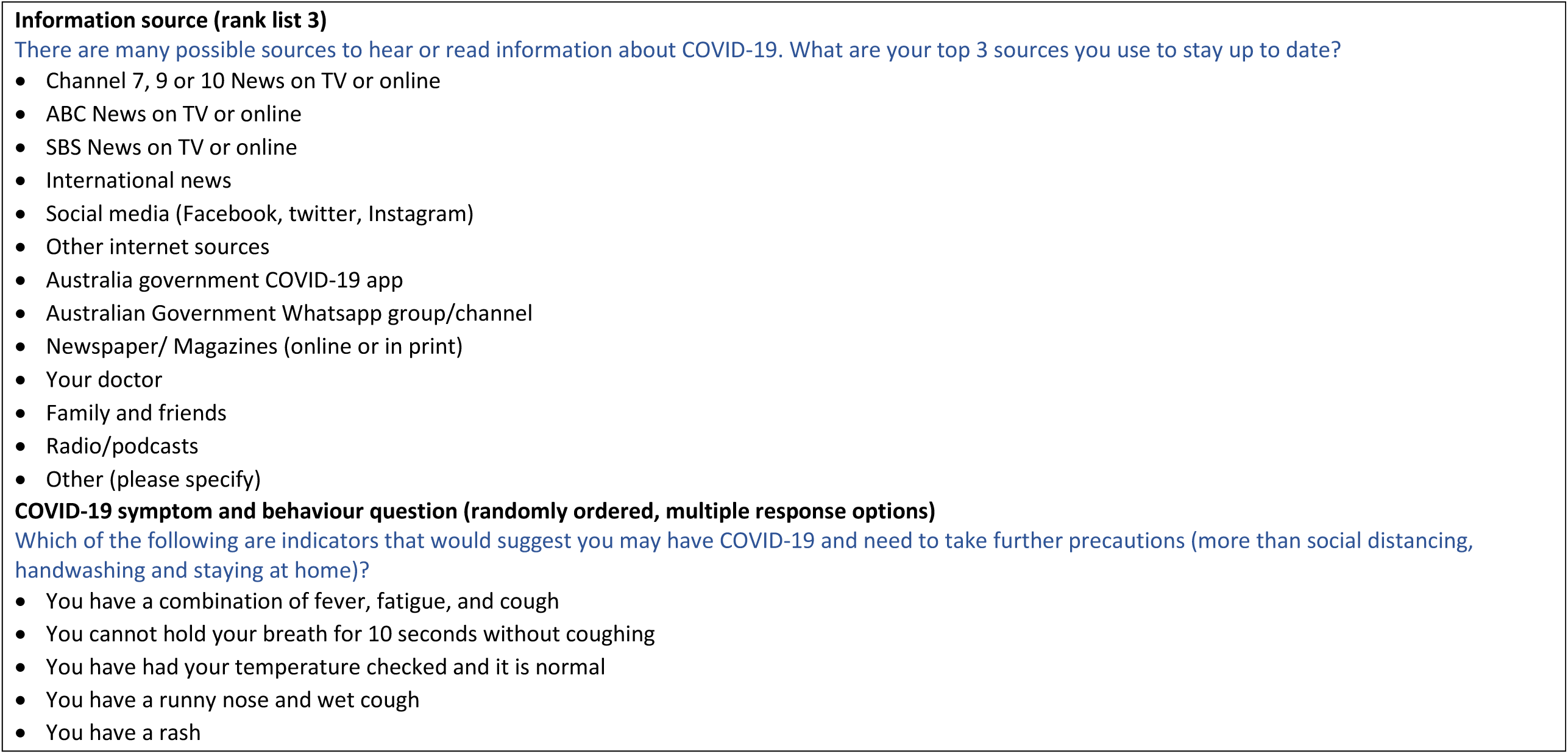
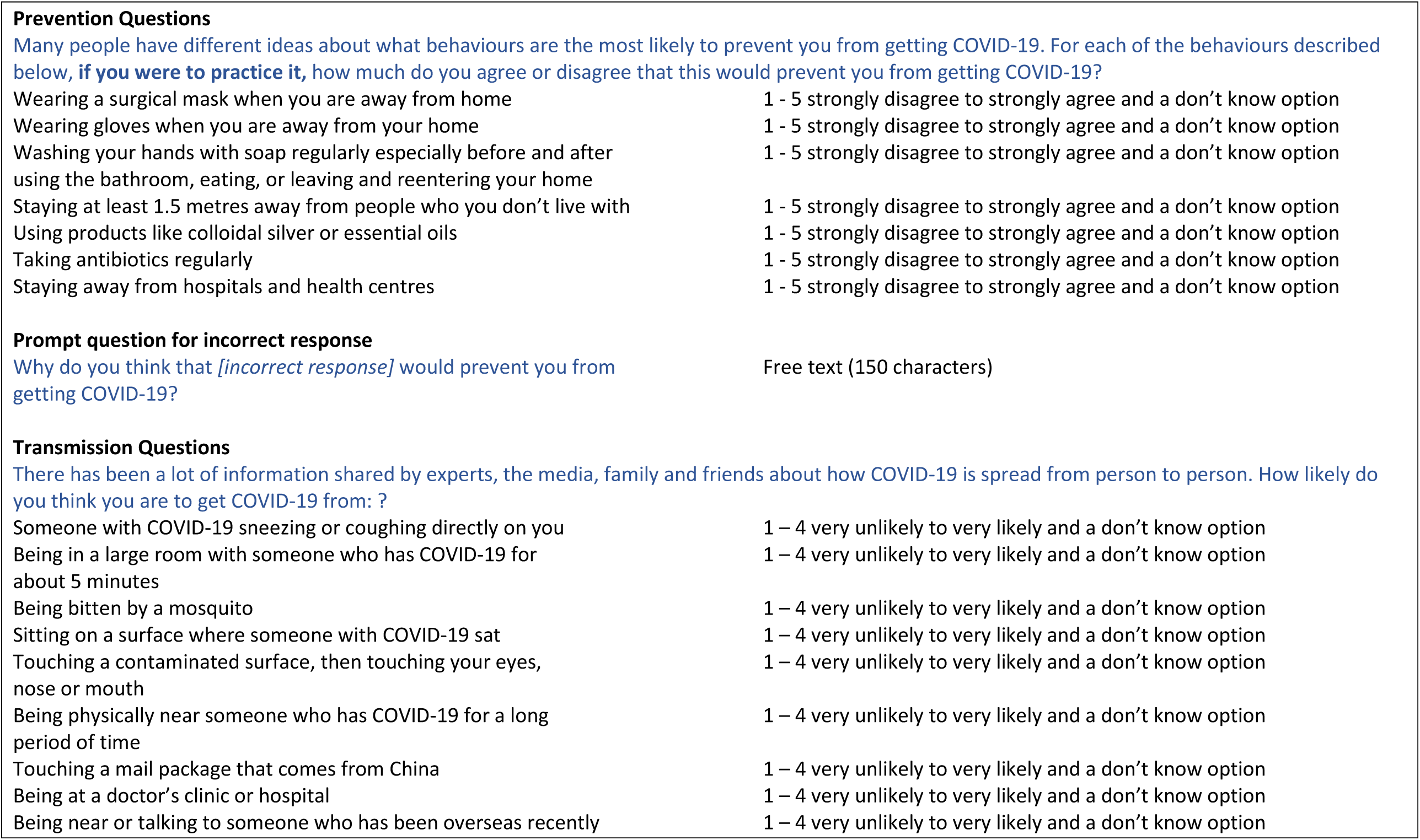
Survey items and scores.

**Supplementary Figure 1.**
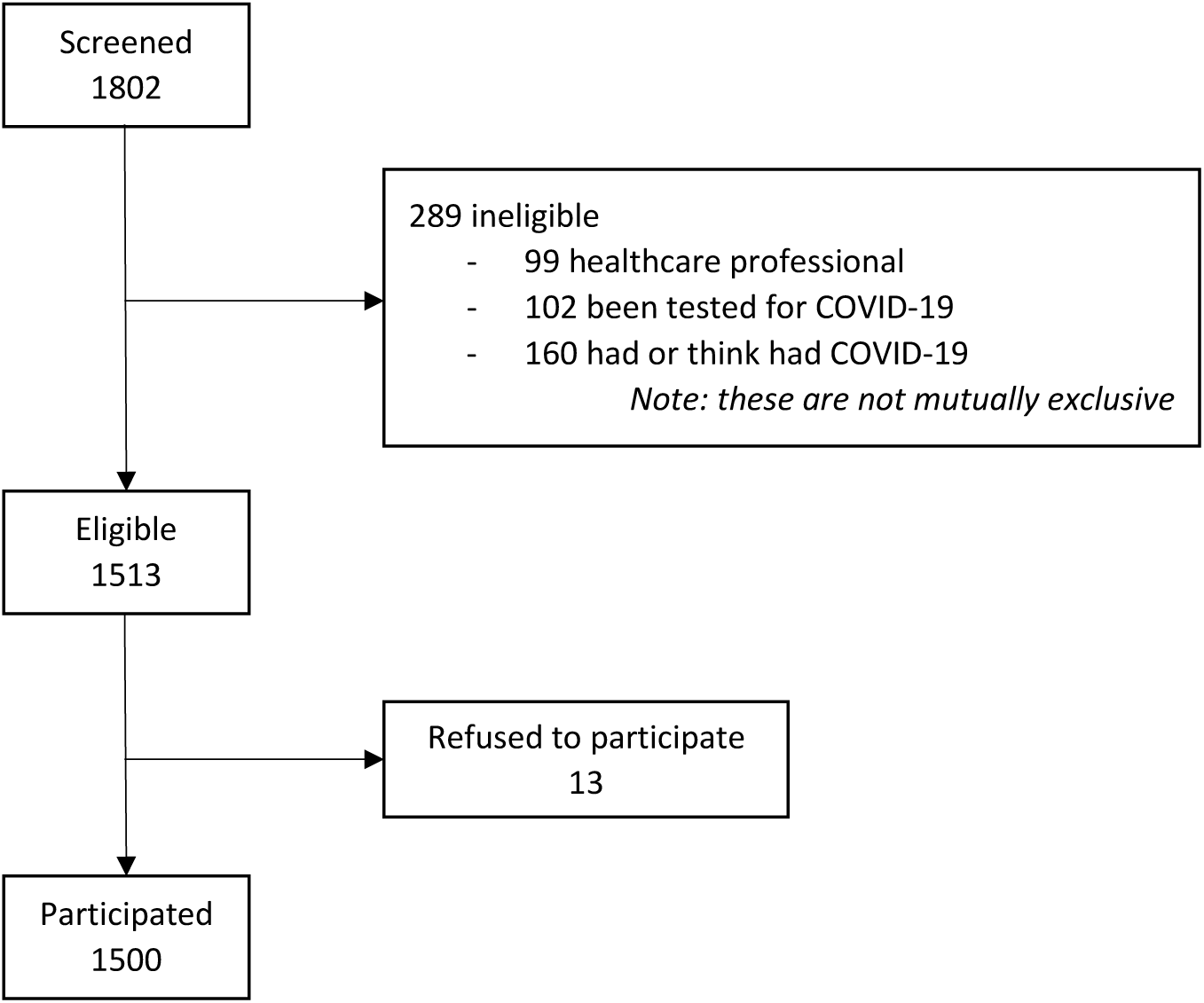
Participant flow chart.

**Supplementary Table 2.** Number (%) of participants’ correct and incorrect responses to COVID-19 symptoms.

| Response | N (%) |
| --- | --- |
| Correct response only | 521 (35) |
| Correct response + 1 incorrect response | 453 (30) |
| Correct response + 2 or more incorrect responses | 369 (25) |
| Incorrect response/s only: | 157 (10) |

**Supplementary Table 3.** Sources of COVID-19 information and their ranking.

| **Sources** | **N**<br>**responses**<br>**(%)**<br>**nominated**<br>**in top 3** | **%**<br>**ranked**<br>**#1** | **%**<br>**ranked**<br>**#2** | **%**<br>**ranked**<br>**#3** |
| --- | --- | --- | --- | --- |
| Channel 7, 9 or 10<br>News on TV or online | 847 (56%) | 48% | 32% | 21% |
| ABC News on TV or<br>online | 647 (43%) | 44% | 32% | 24% |
| Australia government<br>COVID-19 app | 472 (31%) | 44% | 28% | 28% |
| Newspaper/<br>Magazines (online or<br>in print) | 350 (23%) | 29% | 37% | 34% |
| Social media<br>(Facebook, twitter,<br>Instagram) | 342 (23%) | 24% | 40% | 37% |
| Australian<br>Government<br>Whatsapp group | 286 (19%) | 35% | 34% | 31% |
| Family and friends | 260 (17%) | 23% | 34% | 43% |
| Your doctor | 237 (16%) | 24% | 33% | 43% |
| SBS News on TV or<br>online | 236 (16%) | 19% | 42% | 39% |
| International news | 235 (16%) | 23% | 29% | 49% |
| Other internet<br>sources | 235 (16%) | 24% | 32% | 44% |
| Radio/podcasts | 193 (13%) | 17% | 40% | 44% |

Green shading depicts the three highest proportions for ranked information sources

## References

1. Hoffmann T, Glasziou P. What if the vaccine or drugs don’t save us? Plan B for coronavirus means research on alternatives is urgently needed. Online: The Conversation; 2020. Available from: https://theconversation.com/what-if-the-vaccine-or-drugs-dont-save-us-plan-b-for-coronavirus-means-research-on-alternatives-is-urgently-needed-136833 accessed 3rd July

2. Bonell C, Michie S, Reicher S, West R, Bear L, Yardley L, et al. Harnessing behavioural science in public health campaigns to maintain ‘social distancing’ in response to the COVID-19 pandemic: key principles J Epidemiol Community Health 2020;74:617–619.

3. Cane J, O’Connor D, Michie S. Validation of the theoretical domains framework for use in behaviour change and implementation research. Implementation Science. 2012;7(37).

4. Van Bavel JJ, Baicker K, Boggio PS, Capraro V, Cichocka A, Cikara M, et al. Using social and behavioural science to support COVID-19 pandemic response. Nat Hum Behav. 2020;4(5):460–71.

5. McCaffery K, Dodd RH, Cvejic E, Ayre J, Batcup C, Isautier JMJ, et al, Bonner, C. Disparities in COVID-19 related knowledge, attitudes, beliefs and behaviours by health literacy. Public Health Review. 2020, Under Review.

6. Geldsetzer, P. Knowledge and Perceptions of COVID-19 Among the General Public in the United States and the United Kingdom: A Cross-sectional Online Survey. Ann. Intern. Med. 2020;M20-0912 doi: 10.7326/M20-0912

7. Atchison CJ, Bowman L, Vrinten C, Redd R, Pristerà P, Eaton JW, et al. Perceptions and behavioural responses of the general public during the COVID-19 pandemic: A cross-sectional survey of UK Adults medRxiv preprint doi: https://doi.org/10.1101/2020.04.01.20050039.t

8. Seale H, Heywood AE, Leask J, Sheel M, Thomas S, Durrheim DN, et al. COVID-19 is rapidly changing: Examining public perceptions and behaviors in response to this evolving pandemic. PLoS ONE 15(6): e0235112. https://doi.org/10.1371/journal.pone.0235112

9. Wise T, Zbozinek TD, Michelini G, Hagan CC, Mobbs D. Changes in risk perception and protective behavior during the first week of the COVID-19 pandemic in the United States. https://doi.org/10.31234/osf.io/dz428

10. Ventola CL. The Antibiotic Resistance Crisis Part 1: Causes and Threats. P&T, 2015;40(4):3227–283.

11. McCullough AR, Parekh S, Rathbone J, Del mar, CB, Hoffmann T. A systematic review of the public’s knowledge and beliefs about antibiotic resistance. J Antimicrob Chemother 2016; 71: 27–33 doi:10.1093/jac/dkv310

12. Bakhit M, Del Mar C, Gibson E, Hoffmann T. Exploring patients’ understanding of antibiotic resistance and how this may influence attitudes towards antibiotic use for acute respiratory infections: a qualitative study in Australian general practice. BMJ Open 2019;9:e026735. doi:10.1136/bmjopen-2018-026735

13. Medical Benefits Schedule Available from: http://medicarestatistics.humanservices.gov.au/statistics/mbs_item.jsp xaccessed June 10th 2020

14. Hartnett KP, Kite-Powell A, DeVies J, Coletta MA, Boehmer TK, Adjemian J, et al. Impact of the COVID-19 Pandemic on Emergency Department Visits — United States, January 1, 2019–May 30, 2020. Morb. Mortal. Wkly. Rep, 2020;69(23):699–704

15. Lazzerini M, Barbi E, Apicella A, Marchetti F, Cardinale F, Trobia G. Delayed access or provision of care in Italy resulting from fear of COVID-19. Lancet Child Adolesc Health. 2020;4(5):e10–e11. doi:10.1016/S2352-4642(20)30108-5

16. Elshaug A, Duckett S. Hospitals have stopped unnecessary elective surgeries – and shouldn’t restart them after the pandemic. Available from: https://theconversation.com/hospitals-have-stopped-unnecessary-elective-surgeries-and-shouldnt-restart-them-after-the-pandemic-136259 accessed June 17th 2020

